# Exposure to per- and polyfluoroalkyl substances and age-related macular degeneration in U.S. middle-aged and older adults

**DOI:** 10.1101/2024.05.01.24306679

**Authors:** Habyeong Kang, Sung Kyun Park, Dong Hyun Kim, Yoon-Hyeong Choi

## Abstract

Despite various health effects of per- and polyfluoroalkyl substances (PFAS) exposure, the association between PFAS exposure and age-related macular degeneration (AMD) has not been investigated. We aimed to assess associations of PFAS exposure with AMD, using data from 1,722 U.S. adults aged 40 years or more participating in the National Health and Nutrition Examination Survey 2005–2008 with complete data on PFAS measurement, AMD diagnosis, and covariates. Serum concentrations of PFAS, including perfluorooctanoic acid (PFOA), perfluorononanoic acid (PFNA), perfluorohexane sulfonic acid (PFHxS), and perfluorooctane sulfonic acid (PFOS), were measured. An overall PFAS burden score was calculated using item response theory scoring. Individual PFAS concentration and overall PFAS burden score were categorized into low (reference), medium, and high groups. Diagnosis of AMD was based on retinal image examination. Any AMD was defined as the presence of early or late AMD. Survey-weighted logistic regression adjusted for potential confounders was used to calculate odds ratios (ORs) and 95% confidence intervals (CIs) for presence of AMD according to PFAS exposure. Overall, 132 (6.5%) individuals were diagnosed as any AMD, including 115 (5.7%) individuals with early AMD. A significant dose-response association was observed between serum PFOS concentration and any AMD (*p*-trend=0.03), with a significant OR of 1.99 (95% CI: 1.05, 3.79) for the high group compared to the reference. Overall PFAS burden showed a non-monotonic association with any AMD, with a significant OR of 2.18 (95% CI: 1.18, 4.04) for the medium. Inverted U-shaped associations were observed by restricted cubic spline analyses. Also, early AMD showed similar patterns in PFOS and overall PFAS burden and additionally an inverted U-shape association in PFNA. Our findings suggest that exposure to PFAS estimated by serum PFOS and PFNA as well as overall PFAS burden might be a risk factor for AMD in middle-aged and older population.

## 1. Introduction

Age-related macular degeneration (AMD) is one of the leading causes of blindness, accounting for 5.6% of the blindness worldwide (Steinmetz et al., 2021). It has been estimated that the prevalence of early AMD among older adults in the U.S. will gradually increase from 9.1 million in 2010 to 17.8 million in 2050 (Rein, 2009). Therefore, it is important to identify modifiable risk factors for AMD to prevent this disease. In recent years, several studies have reported associations between AMD and environmental factors, such as exposure to ambient air pollution (Ju et al., 2022) and heavy metals (Park et al., 2015; Wu et al., 2014). However, it remains unclear if AMD is associated with exposure to per- and polyfluoroalkyl substances (PFAS), another group of persistent and universal environmental contaminants.

PFAS are man-made chemicals widely applied to various industrial and consumer products, such as non-stick cookware, water-repellent textiles, food packaging, cleaning and degreasing solvents, and firefighting foam agents (Christensen and Calkins, 2023; Glüge et al., 2020). Long-chain PFAS, so-called “forever chemicals”, do not break down. They will persist in the environment and accumulate in the food chain due to their long-lasting chemical properties (Joudan and Lundgren, 2022). PFAS have been detected in ubiquitous environmental matrices of surface water, groundwater, drinking water, soil, wildlife, fish, and shellfish around the world (ATSDR, 2021; Domingo and Nadal, 2019; McCarthy et al., 2017; Ruffle et al., 2020). PFAS exposure in the general population can occur through multiple routes, including ingestion of contaminated foods and/or drinking water and direct contact with PFAS-containing consumer products and ocular device (Domingo and Nadal, 2019; Glüge et al., 2020; Kang et al., 2024; Ragnarsdóttir et al., 2022). Indeed, detectable levels of PFAS exposure biomarkers in serum samples from the general population have been consistently reported worldwide including the U.S. (Calafat et al., 2019), Europe (Bil et al., 2023), and Asia (Seo et al., 2018; Tsai et al., 2018; Zeeshan et al., 2020). Growing epidemiological evidence suggests possible associations between PFAS exposure and various health outcomes, including liver damage (Costello et al., 2022), hyperlipidemia (ATSDR, 2021; Winquist and Steenland, 2014), diabetes (Gui et al., 2023), and reproductive disorders (ATSDR, 2021; Ding et al., 2020), indicating underlying mechanisms including oxidative stress and metabolic and endocrine disruption.

Despite the widespread exposure to PFAS and plausible biological mechanisms, adverse health effects of PFAS on the retina remain unanswered. Thus, the objective of this study was to examine the association between serum PFAS concentrations and AMD in a representative sample of U.S. middle-aged and older adults from the National Health and Nutrition Examination Survey (NHANES).

## 2. Methods

### 2.1. Study population and data collection

The NHANES is an on-going survey of the nationally representative U.S. population conducted by the National Center for Health Statistics (NCHS) of the Centers for Disease Control and Prevention (CDC) (CDC, 2012). The NHANES collects health and nutrition-related data from participants through standardized survey interview, medical examination, biospecimen collection, and laboratory analysis (CDC, 2012). We used data of adults aged 40 years or older who participated in the NHANES cycles of 2005–2006 and 2007–2008 because ophthalmology examinations were conducted only in these two cycles. In NHANES 2005–2008, serum PFAS concentrations were measured in a one-third random sample of eligible participants. For this analysis, we included participants aged 40 years or older with complete data on retinal image examination, PFAS measurements, and covariates, resulting in a final sample of 1,722 participants (Supplemental Fig. S1). The NHANES study protocol was approved by the NCHS Research Ethics Review Board. All participants provided informed consent before participating in the study. This study was conducted following the Declaration of Helsinki.

### 2.2. Assessment of PFAS exposure

PFAS exposure was assessed by measuring serum PFAS concentrations. Analytical methods were described in a previous study (Kuklenyik et al., 2005). Briefly, serum was diluted with formic acid and PFAS were preconcentrated using an online solid-phase extraction. Individual PFAS were then separated and quantified using a high-performance liquid chromatography−turbo ion spray ionization-tandem mass spectrometry (HPLC-TIS-MS/MS). Target PFAS included perfluorheptanoic acid (PFHpA), perfluorooctanoic acid (PFOA), perfluorononanoic acid (PFNA), perfluorodecanoic acid (PFDA), perfluoroundecanoic acid (PFUnDA), perflurododecanoic acid (PFDoDA), perfluorobutane sulfonic acid (PFBS), perfluorohexane sulfonic acid (PFHxS), perfluorooctane sulfonic acid (PFOS), 2-(N-Methyl-perfluorooctane sulfonamido) acetic acid (MeFOSAA), 2-(N-Ethyl-perfluorooctane sulfonamido) acetic acid (EtFOSAA), and perfluorooctane sulfonamide (PFOSA). Limits of detection (LOD) ranged from 0.082 to 0.4 ng/mL depending on the compound (Supplemental Table S1). Concentrations below LOD were imputed using LOD divided by square root of 2.

We estimated an overall PFAS burden score for each participant using the item response theory (IRT) (Liu et al., 2022). IRT was chosen because individual PFAS compounds share common exposure sources and similar toxicological mechanisms (Jane L Espartero et al., 2022; Liu et al., 2022). IRT is a well-established latent variable method used in education research for test scoring. It has been recently adopted for environmental exposure burden scoring such as PFAS (Liu et al., 2022). IRT can estimate participants’ latent PFAS burden based on their concentrations of individual PFAS and their exposure patterns, thereby allowing us to compare overall burden of PFAS exposure across participants. We used a graded response IRT model to compute IRT score for overall PFAS burden following the method previously described by Liu et al. (2022) of “factor.scores” in R package “Itm’ (version 1.2-0). In IRT scoring, we included PFOA, PFNA, PFDA, PFUnDA, PFHxS, PFOS, and MeFOSAA whereas we did not consider PFHpA, PFDoDA, PFBS, EtFOSAA, and PFOSA in the IRT scoring due to their low detection frequencies (< 20%). Those with detection frequencies over 90% (i.e., PFOA, PFNA, PFHxS, PFOS) were categorized into deciles, while MeFOSAA, PFDA, and PFUnDA were categorized into quartile, tertile, and binary depending on their detection frequencies (75.0%, 71.7%, and 36.1%, respectively).

### 2.3. Diagnosis of age-related macular disease

Retinal images of participants were captured using a retinal camera (CR6-45NM Non-Mydriatic Retinal Camera; Canon, Inc., Tokyo, Japan). Details of camera settings were described elsewhere (CDC, 2005). Images were then graded by trained ophthalmic graders based on a modified method of the Wisconsin Age-Related Maculopathy Grading System (Klein et al., 1991). If the disease severity was assessed in both eyes, results from the worse eye were used. Participants who had any late lesions such as exudative AMD signs or geographic atrophy in at least one eye were diagnosed as late AMD. Among those without late AMD signs, participants were diagnosed as early AMD if the following signs were observed in at least one eye: 1) presence of soft drusen with a grid area of greater than a 500 µ circle and a pigmentary abnormality, 2) presence of soft drusen in the center circle and a pigmentary abnormality. Any AMD was defined as the presence of early or late AMD.

### 2.4. Covariates

Covariates were chosen based on prior knowledge of relationships between covariates and PFAS exposure as well as relationships between covariates and the development of AMD. We considered sex, race/ethnicity, age, education, smoking history, body mass index (BMI), seafood consumption, and estimated glomerular filtration rate (eGFR). Race/ethnicity was categorized as Non-Hispanic White, Non-Hispanic Black, Hispanic, or other race/ethnicity based on self-identification during survey interview. Age was considered both linear and squared terms considering a potential non-linear relationship between age and AMD risk (Ju et al., 2022; Wu et al., 2014). Education was categorized as ‘less than high school’, ‘high school diploma’, or ‘some college or more’. Cumulative smoking pack-year was categorized as non-smoker, light smoker (> 0 to < 20 pack-years), or heavy smoker (≥ 20 pack-years). BMI (kg/m^2^) was calculated as measured weight (kg) divided by the square of measured height (m). Dietary seafood consumption was calculated as a sum of the number of times each fish or shellfish item was eaten in the last 30 days, which was self-reported by each participant. We calculated eGFR using the CKD-EPI 2021 equation (Inker et al., 2021).

### 2.5. Statistical Analysis

All statistical analyses were conducted using R version 4.2.2. Statistical significance was considered when two-sided *p-*value was less than 0.05. The complex sample design and sampling weights of the NHANES data were accounted for using the R package “survey” (version 4.1-1).

Associations of PFAS with AMD were evaluated using survey-weighted logistic regression. Given the small proportion of late AMD cases, any AMD was considered the primary outcome measure and results of early AMD were reported as a secondary outcome measure. PFAS levels of individual compounds and their IRT score were modeled as tertile groups (denoted ‘low’, ‘medium’, and ‘high’, respectively). We assessed the models with different sets of covariates. Model 1 was adjusted for core demographic variables, including sex, race/ethnicity, age, and age square. Model 2, which was considered the main model in this study, was further adjusted for education, smoking pack-year, BMI, and seafood consumption. Model 3 was additionally adjusted for eGFR to account for potential effects of kidney function on excretion of PFAS into urine (Jain and Ducatman, 2019; Park et al., 2021).

Non-linear associations of PFAS concentrations with AMD were visualized using restricted cubic splines with survey-weighted logistic regression. They were created using the function “ns” in the R package “splines” (version 4.2.2). Two knots were located at the 33^rd^ and 67^th^ percentiles of each PFAS concentration.

A subgroup analysis was conducted for the association between overall PFAS burden and any AMD in subgroups divided by age (< 60 or ≥ 60 years), sex (male or female), race/ethnicity (Non-Hispanic White or others), or seafood consumption (< 4 or ≥ 4 times per month).

Also, in order to ensure robustness of associations, we conducted a sensitivity analysis with additional adjustment for cardiometabolic status (i.e., hypertension, diabetes, and/or total cholesterol level) and blood cadmium concentrations as potential risk factors affecting AMD (Gemmy Cheung et al., 2013; Ghaem Maralani et al., 2015; Wu et al., 2014). We defined hypertension as self-report of diagnosis, use of medications for high blood pressure, diastolic blood pressure ≥ 90 mm Hg, or systolic blood pressure ≥ 140 mm Hg. Diabetes was defined as self-report of diagnosis or the use of insulin or oral hypoglycemic medications.

## 3. Results

### 3.1. Demographic characteristics of participants by AMD status

Of the 1,722 participants, 132 were diagnosed with AMD including 115 with early AMD and 17 with late AMD. Participants with AMD were more likely to be older and Non-Hispanic White. They were more likely to have diabetes than those without AMD (Table 1). Additionally, participants with AMD had lower eGFR and higher concentrations of PFHxS and PFOS than those without AMD. Among PFAS measured in serum, PFOS had the highest concentration, followed by PFOA, PHFxS, and PFNA.

**Table 1.** Survey-weighted participants’ characteristics by age-related macular degeneration (AMD) status among U.S. adults aged 40 years or older participating in the NHANES 2005–2008.

|  |  | n (weighted %) or weighted mean±SE |  |  | p-value<sup>a</sup> |
| --- | --- | --- | --- | --- | --- |
|  |  | Total<br>(n=1722) | Non-AMD<br>(n=1590) | Any AMD<br>(n=132) |  |
| Early AMD |  | 115 (5.7) |  | 115 (87.4) |  |
| Late AMD |  | 17 (0.8) |  | 17 (12.6) |  |
| Age (year) |  | 57.0±0.4 | 56.1±0.4 | 69.7±1.6 | <0.0001 |
| Female |  | 843 (52.0) | 784 (52.3) | 59 (47.9) | 0.37 |
| Race/ethnicity | Non-Hispanic White | 950 (77.8) | 852 (77.0) | 98 (89.3) | 0.0003 |
|  | Non-Hispanic Black | 323 (8.9) | 314 (9.4) | 9 (2.2) |  |
|  | Hispanic | 384 (8.4) | 362 (8.5) | 22 (6.9) |  |
|  | Other | 65 (4.9) | 62 (5.2) | 3 (1.6) |  |
| Education | Less than high school | 501 (18.7) | 467 (18.6) | 34 (20.2) | 0.90 |
|  | High school diploma | 409 (24.9) | 375 (24.9) | 34 (24.4) |  |
|  | Some college+ | 812 (56.4) | 748 (56.5) | 64 (55.4) |  |
| Cumulative<br>smoking pack-year | Non-smoker | 828 (49.9) | 774 (50.6) | 54 (40.3) | 0.15 |
|  | Light smoker | 476 (24.9) | 438 (24.7) | 38 (27.7) |  |
|  | Heavy smoker | 418 (25.2) | 378 (24.7) | 40 (32.1) |  |
| Body mass index (kg/m<sup>2</sup>) |  | 29±0.2 | 29±0.3 | 28.3±0.5 | 0.22 |
| Hypertension<sup>b</sup> |  | 823 (42.2) | 746 (41.3) | 77 (54.7) | 0.50 |
| Diabetes<sup>b</sup> |  | 260 (11.6) | 243 (11.5) | 17 (13.8) | 0.03 |
| Total cholesterol (mg/dL)<sup>b</sup> |  | 205.5±1.1 | 205.5±1.1 | 206.5±4.4 | 0.82 |
| eGFR (mL/min/1.73 m<sup>2</sup>)<sup>c</sup> |  | 86.6±0.8 | 87.3±0.8 | 77.2±2.4 | 0.0002 |
| Seafood<br>consumption in<br>past 30 days | <3 times | 722 (38.1) | 665 (37.9) | 57 (41.1) | 0.30 |
|  | 3-5 times | 481 (28.4) | 446 (28.9) | 35 (21.8) |  |
|  | >5 times | 519 (33.5) | 479 (33.2) | 40 (37.1) |  |
| PFOA (ng/mL)<sup>d</sup> |  | 4.3±1.0 | 4.3±1.0 | 4.7±1.1 | 0.11 |
| PFNA (ng/mL)<sup>d</sup> |  | 1.2±1.1 | 1.2±1.1 | 1.3±1.1 | 0.32 |
| PFHxS (ng/mL)<sup>d</sup> |  | 1.9±1.1 | 1.9±1.1 | 2.2±1.1 | 0.04 |
| PFOS (ng/mL)<sup>d</sup> |  | 17.2±1.0 | 16.9±1.0 | 21.9±1.1 | 0.0004 |
| Overall PFAS burden (IRT score)<sup>e</sup> |  | 0.00±0.06 | -0.01±0.06 | 0.15±0.09 | 0.10 |

Note: n (%) for categorical variables; weighted mean±SE (standard error). NHANES, National Health and Nutrition Examination Survey; eGFR, estimated glomerular filtration rate; PFOA, perfluorooctanoic acid; PFNA, perfluorononanoic acid; PFHxS, perfluorohexane sulfonic acid; PFOS, perfluorooctane sulfonic acid; PFAS, per- and polyfluoroalkyl substances; IRT, item response theory.
<sup>a</sup> Survey-weighted chi-square test for categorical variables and survey-weighted t-test for continuous variables.
<sup>b</sup> n=1702.
<sup>c</sup> Estimated glomerular filtration rate, n=1718.
<sup>d</sup> Geometric mean±geometric standard error.
<sup>e</sup> Overall PFAS burden was derived by IRT scoring<sup>26</sup> using serum concentrations of PFOA, PFNA, PFDA, PFUnDA, PFHxS, PFOS, and MeFOSAA.

### 3.2. Overall PFAS burden and correlation with individual PFAS compounds

Item information curve showed that PFNA highly contributed to overall PFAS burden (IRT score), which was indicated by a higher level of its information curve than other PFAS compounds (Supplemental Fig. S2). Individual PFAS compounds were observed to be correlated with each other (Spearman’s correlation coefficients (r): 0.07 to 0.81), and overall PFAS burden (IRT score) showed high correlations with individual PFAS compounds, particularly with PFNA (r = 0.94) and PFDA (r = 0.88) (Supplemental Fig. S3 and Table S2).

### 3.3. Associations of serum PFAS with AMD

Serum concentration of PFOS was positively associated with any AMD, exhibiting an OR of 1.99 (95% CI: 1.05, 3.79) for the high group compared to the low group (reference), along with a significant linear trend (*p* = 0.03 in Model 2; Table 2). While the association of PFNA with any AMD was not monotonous, a positive association was observed for the medium group with marginal significance, showing an OR of 1.74 (95% CI: 0.98, 3.09). Overall PFAS burden also had a non-monotonous association with any AMD (OR: 2.18, 95% CI: 1.18, 4.04 for the medium group; OR: 1.88, 95% CI: 0.96, 3.68 for the high group). Associations between PFAS and any AMD were consistent across models with different sets of covariates (Model 1 and Model 3).

**Table 2.**
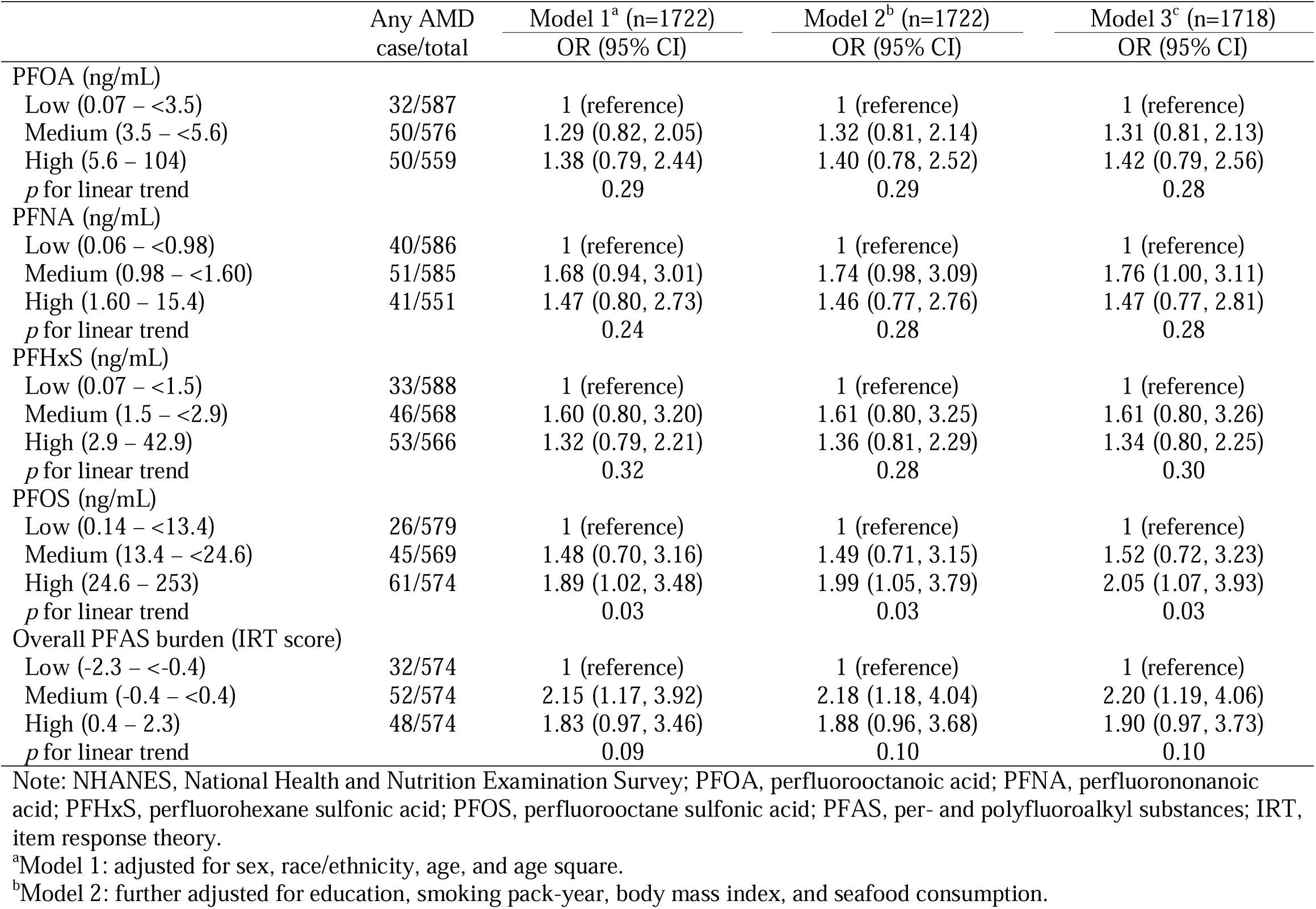
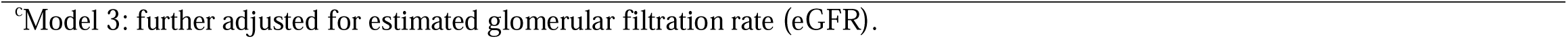
Survey-weighted odds ratios (ORs) and 95% confidence intervals (CIs) for any age-related macular degeneration (AMD) by serum PFAS levels among U.S. adults aged 40 years or older participating in the NHANES 2005–2008.

|  | Any AMD<br>case/total | Model 1<sup>a</sup> (n=1722)<br>OR (95% CI) | Model 2<sup>b</sup> (n=1722)<br>OR (95% CI) | Model 3<sup>c</sup> (n=1718)<br>OR (95% CI) |
| --- | --- | --- | --- | --- |
| PFOA (ng/mL) |  |  |  |  |
| Low (0.07 – <3.5) | 32/587 | 1 (reference) | 1 (reference) | 1 (reference) |
| Medium (3.5 – <5.6) | 50/576 | 1.29 (0.82, 2.05) | 1.32 (0.81, 2.14) | 1.31 (0.81, 2.13) |
| High (5.6 – 104) | 50/559 | 1.38 (0.79, 2.44) | 1.40 (0.78, 2.52) | 1.42 (0.79, 2.56) |
| *p* for linear trend |  | 0.29 | 0.29 | 0.28 |
| PFNA (ng/mL) |  |  |  |  |
| Low (0.06 – <0.98) | 40/586 | 1 (reference) | 1 (reference) | 1 (reference) |
| Medium (0.98 – <1.60) | 51/585 | 1.68 (0.94, 3.01) | 1.74 (0.98, 3.09) | 1.76 (1.00, 3.11) |
| High (1.60 – 15.4) | 41/551 | 1.47 (0.80, 2.73) | 1.46 (0.77, 2.76) | 1.47 (0.77, 2.81) |
| *p* for linear trend |  | 0.24 | 0.28 | 0.28 |
| PFHxS (ng/mL) |  |  |  |  |
| Low (0.07 – <1.5) | 33/588 | 1 (reference) | 1 (reference) | 1 (reference) |
| Medium (1.5 – <2.9) | 46/568 | 1.60 (0.80, 3.20) | 1.61 (0.80, 3.25) | 1.61 (0.80, 3.26) |
| High (2.9 – 42.9) | 53/566 | 1.32 (0.79, 2.21) | 1.36 (0.81, 2.29) | 1.34 (0.80, 2.25) |
| *p* for linear trend |  | 0.32 | 0.28 | 0.30 |
| PFOS (ng/mL) |  |  |  |  |
| Low (0.14 – <13.4) | 26/579 | 1 (reference) | 1 (reference) | 1 (reference) |
| Medium (13.4 – <24.6) | 45/569 | 1.48 (0.70, 3.16) | 1.49 (0.71, 3.15) | 1.52 (0.72, 3.23) |
| High (24.6 – 253) | 61/574 | 1.89 (1.02, 3.48) | 1.99 (1.05, 3.79) | 2.05 (1.07, 3.93) |
| *p* for linear trend |  | 0.03 | 0.03 | 0.03 |
| Overall PFAS burden (IRT score) |  |  |  |  |
| Low (-2.3 – <-0.4) | 32/574 | 1 (reference) | 1 (reference) | 1 (reference) |
| Medium (-0.4 – <0.4) | 52/574 | 2.15 (1.17, 3.92) | 2.18 (1.18, 4.04) | 2.20 (1.19, 4.06) |
| High (0.4 – 2.3) | 48/574 | 1.83 (0.97, 3.46) | 1.88 (0.96, 3.68) | 1.90 (0.97, 3.73) |
| *p* for linear trend |  | 0.09 | 0.10 | 0.10 |

Note: NHANES, National Health and Nutrition Examination Survey; PFOA, perfluorooctanoic acid; PFNA, perfluorononanoic acid; PFHxS, perfluorohexane sulfonic acid; PFOS, perfluorooctane sulfonic acid; PFAS, per- and polyfluoroalkyl substances; IRT, item response theory.
<sup>a</sup>Model 1: adjusted for sex, race/ethnicity, age, and age square.
<sup>b</sup>Model 2: further adjusted for education, smoking pack-year, body mass index, and seafood consumption.
<sup>c</sup>Model 3: further adjusted for estimated glomerular filtration rate (eGFR).

Trends for associations of PFAS with early AMD were generally similar to those observed for associations of PFAS with any AMD, but they were stronger than those observed for any AMD (Table 3). Specifically, in Model 2, OR was 2.03 (95% CI: 1.17, 3.51) for the medium group of PFNA and 2.45 (95% CI: 1.30, 4.63) for the overall PFAS burden.

**Table 3.**
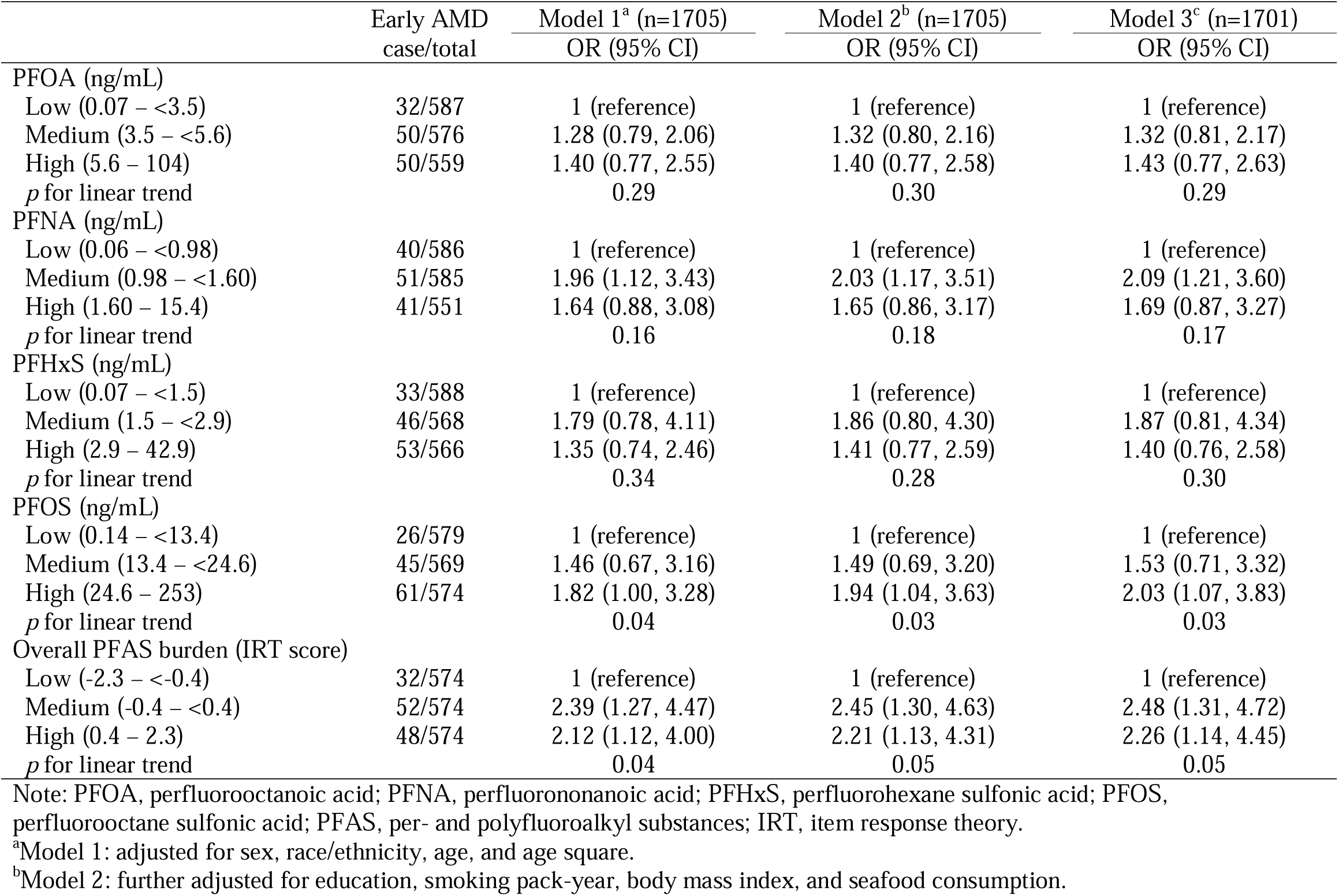
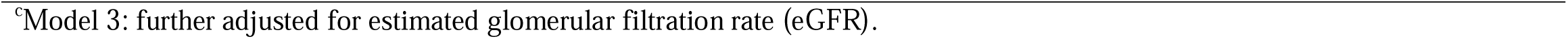
Survey-weighted odds ratios (ORs) and 95% confidence intervals (CIs) for early age-related macular degeneration (AMD) by serum PFAS levels in subpopulation excluding subjects with late AMD.

|  | Early AMD<br>case/total | Model 1<sup>a</sup> (n=1705)<br>OR (95% CI) | Model 2<sup>b</sup> (n=1705)<br>OR (95% CI) | Model 3<sup>c</sup> (n=1701)<br>OR (95% CI) |
| --- | --- | --- | --- | --- |
| PFOA (ng/mL) |  |  |  |  |
| Low (0.07 – <3.5) | 32/587 | 1 (reference) | 1 (reference) | 1 (reference) |
| Medium (3.5 – <5.6) | 50/576 | 1.28 (0.79, 2.06) | 1.32 (0.80, 2.16) | 1.32 (0.81, 2.17) |
| High (5.6 – 104) | 50/559 | 1.40 (0.77, 2.55) | 1.40 (0.77, 2.58) | 1.43 (0.77, 2.63) |
| *p* for linear trend |  | 0.29 | 0.30 | 0.29 |
| PFNA (ng/mL) |  |  |  |  |
| Low (0.06 – <0.98) | 40/586 | 1 (reference) | 1 (reference) | 1 (reference) |
| Medium (0.98 – <1.60) | 51/585 | 1.96 (1.12, 3.43) | 2.03 (1.17, 3.51) | 2.09 (1.21, 3.60) |
| High (1.60 – 15.4) | 41/551 | 1.64 (0.88, 3.08) | 1.65 (0.86, 3.17) | 1.69 (0.87, 3.27) |
| *p* for linear trend |  | 0.16 | 0.18 | 0.17 |
| PFHxS (ng/mL) |  |  |  |  |
| Low (0.07 – <1.5) | 33/588 | 1 (reference) | 1 (reference) | 1 (reference) |
| Medium (1.5 – <2.9) | 46/568 | 1.79 (0.78, 4.11) | 1.86 (0.80, 4.30) | 1.87 (0.81, 4.34) |
| High (2.9 – 42.9) | 53/566 | 1.35 (0.74, 2.46) | 1.41 (0.77, 2.59) | 1.40 (0.76, 2.58) |
| *p* for linear trend |  | 0.34 | 0.28 | 0.30 |
| PFOS (ng/mL) |  |  |  |  |
| Low (0.14 – <13.4) | 26/579 | 1 (reference) | 1 (reference) | 1 (reference) |
| Medium (13.4 – <24.6) | 45/569 | 1.46 (0.67, 3.16) | 1.49 (0.69, 3.20) | 1.53 (0.71, 3.32) |
| High (24.6 – 253) | 61/574 | 1.82 (1.00, 3.28) | 1.94 (1.04, 3.63) | 2.03 (1.07, 3.83) |
| *p* for linear trend |  | 0.04 | 0.03 | 0.03 |
| Overall PFAS burden (IRT score) |  |  |  |  |
| Low (-2.3 – <-0.4) | 32/574 | 1 (reference) | 1 (reference) | 1 (reference) |
| Medium (-0.4 – <0.4) | 52/574 | 2.39 (1.27, 4.47) | 2.45 (1.30, 4.63) | 2.48 (1.31, 4.72) |
| High (0.4 – 2.3) | 48/574 | 2.12 (1.12, 4.00) | 2.21 (1.13, 4.31) | 2.26 (1.14, 4.45) |
| *p* for linear trend |  | 0.04 | 0.05 | 0.05 |

Note: PFOA, perfluorooctanoic acid; PFNA, perfluorononanoic acid; PFHxS, perfluorohexane sulfonic acid; PFOS, perfluorooctane sulfonic acid; PFAS, per- and polyfluoroalkyl substances; IRT, item response theory.
<sup>a</sup>Model 1: adjusted for sex, race/ethnicity, age, and age square.
<sup>b</sup>Model 2: further adjusted for education, smoking pack-year, body mass index, and seafood consumption.
<sup>c</sup>Model 3: further adjusted for estimated glomerular filtration rate (eGFR).

### 3.4. Non-linear association of serum PFAS with AMD

PFNA, PFOS, and overall PFAS burden, which had shown significant results in linear regressions (Tables 2 and 3), were examined using restricted cubic spline curves to evaluate non-linearity (Fig. 1). Curves for PFNA, PFOS, and overall PFAS burden showed an inverted U-shaped association. Concentration-response shapes were positive for PFNA < ~1.7 ng/mL, PFOS < ~31.2 ng/mL, and overall PFAS burden < ~0.61. However, at concentrations higher than those thresholds, curves showed a negative association between serum PFAS and any AMD or early AMD.

**Figure 1.**
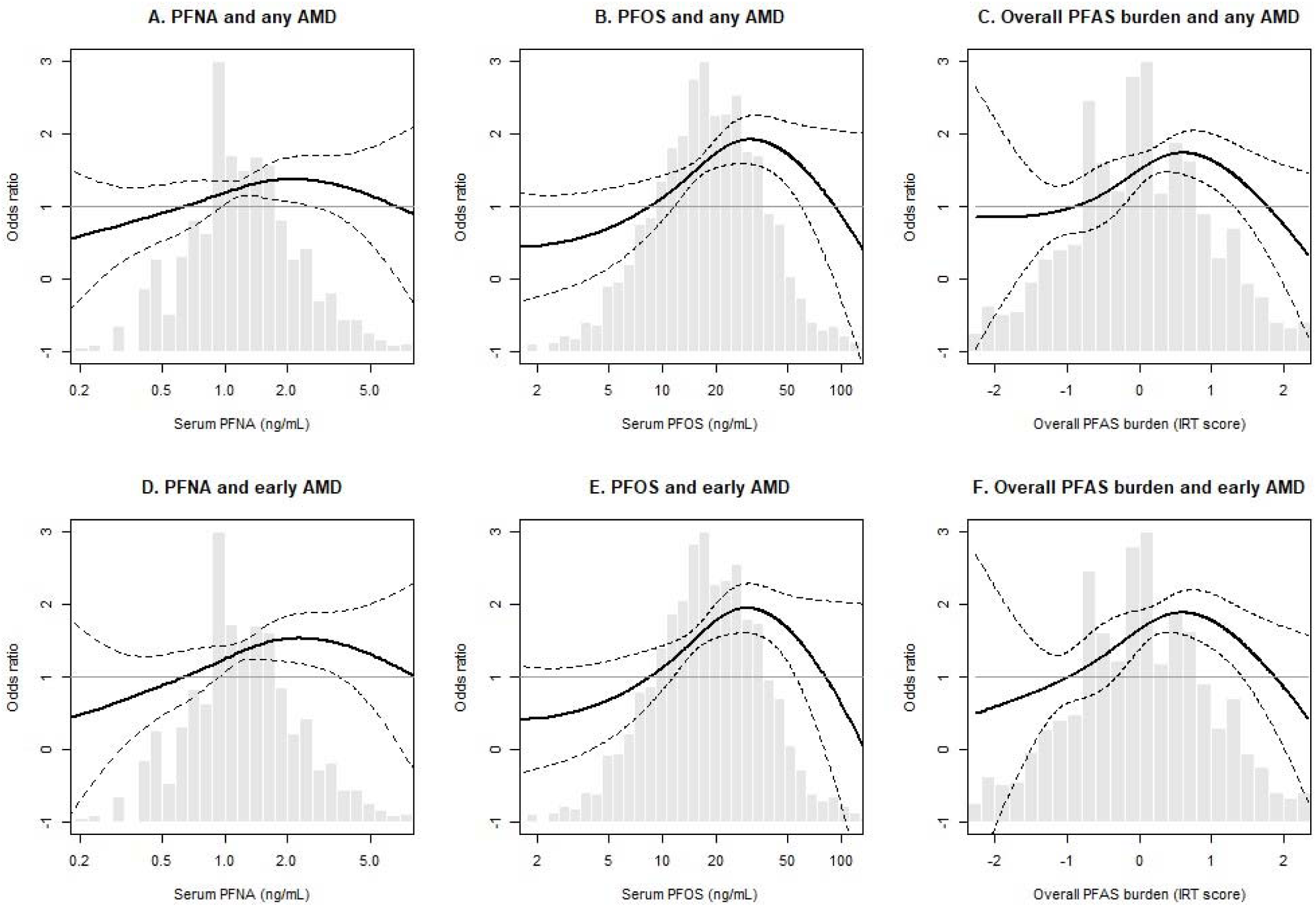
Non-linear associations of serum PFAS concentrations with any or early age-related macular degeneration (AMD) among U.S. adults aged 40 years or older participating in the National Health and Nutrition Examination Survey (NHANES) 2005–2008 (n = 1,722 for any AMD and n = 1,705 for early AMD). Restricted cubic spline with survey-weighted logistic regression adjusted for sex, race/ethnicity, age, age square, education, smoking pack-year, body mass index, and seafood consumption was used. Two knots were located at 33^rd^ and 67^th^ percentiles. Adjusted odds ratios (ORs) and their 95% confidence intervals are presented as bold lines and dotted lines, respectively. First tertile group of each per- and polyfluoroalkyl substances (PFAS) concentration was set as the reference (average OR of reference = 1). Histograms of PFAS concentrations are presented as gray bars. Note: PFNA, perfluorononanoic acid; PFOS, perfluorooctane sulfonic acid.

### 3.5. Subgroup analysis

The association of overall PFAS burden with any AMD was evaluated for different subgroups (Fig. 2). A stronger association was observed for middle-aged adults of 40-59 years compared to older adults ≥ 60 years. Additionally, the association was stronger for males than for females and for Non-Hispanic White participants than for other race/ethnicity. Participants with high seafood consumption (≥ 4 times/month) showed a greater association than for those with lower seafood consumption (< 4 times/month).

**Figure 2.**
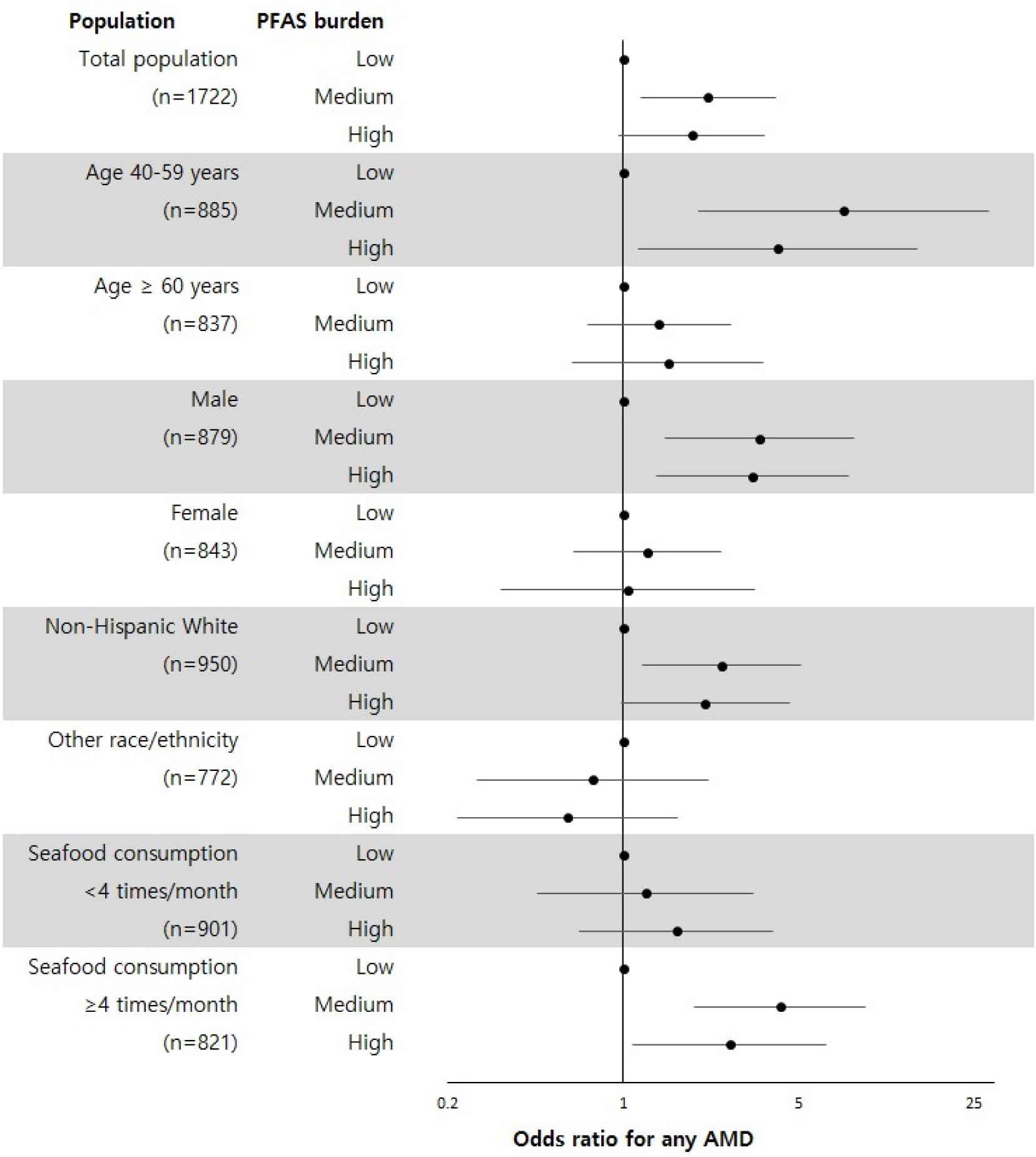
Subgroup analysis for the association of overall PFAS burden with any age-related macular degeneration (AMD) among U.S. adults aged 40 years or older participating in the National Health and Nutrition Examination Survey (NHANES) 2005–2008 (n = 1,722). Logistic regression models were adjusted for sex, race/ethnicity, age, age square, education, smoking pack-year, body mass index, and seafood consumption. Low per- and polyfluoroalkyl substances (PFAS) burden was set as the reference. Odds ratios (ORs) are presented as dots and their 95% confidence intervals (CIs) are presented as bars.

### 3.6. Sensitivity analysis

To assess the robustness of results, we conducted a sensitivity analysis by further adjusting for hypertension, diabetes, and/or total cholesterol (Supplemental Table S3), and blood cadmium concentrations (Supplemental Table S4). We observed that the additional adjustment did not alter observed associations between PFAS and any AMD.

## 4. Discussion

To the best of our knowledge, this is the first epidemiological study to investigate the potential association between PFAS exposure and AMD. In the present study using U.S. middle-aged and older adults from the NHANES 2005-2008, any AMD had a dose-dependent association with an increased serum PFOS and a non-monotonic association with an increased overall PFAS burden. Additionally, early AMD had a non-monotonic association with an increased serum PFNA, which provided compelling evidence that PFAS exposure could be linked to an increased risk of AMD even at exposure levels observed in the general population.

It is difficult to compare our findings to previous evidence because limited research is available on associations between serum PFAS concentrations and ocular health outcomes such as AMD. Only one recent study in China has investigated cross-sectional associations of serum PFAS concentrations and ocular conditions including macular disorder in industrial city dwellers (n = 1202) with high levels of PFAS exposure (Zeeshan et al., 2020). Although that study evaluated various linear and branched PFAS, only perfluorododecanoic acid (PFDoDA) concentration showed a significant association with macular disorder. However, significant associations of PFOS and PFNA observed in our study were not observed in Chinese industrial dwellers. These different findings might be due to different PFAS exposure levels and outcome measures between the two studies. The current study included a general population with low-to-high range of PFAS exposure, while the Chinese study included only highly exposed population in the city near fluoropolymer manufacturing facilities (Zeeshan et al., 2020). Also, the current study used a globally recognized measure for AMD diagnosis, i.e., the Wisconsin Age-Related Maculopathy Grading System, while the Chinese study used an ocular condition for macular disorder, i.e., pigment disorder in the macular area of the eye (Zeeshan et al., 2020). Thus, differences in results might be due to differences in study design rather than different etiology.

Several hypotheses can be suggested as potential biological mechanisms linking PFAS exposure to the risk of AMD. Oxidative stress is one of the potential biological mechanisms. Oxidative stress is known to be one of the key pathways in AMD development (Armento et al., 2021). In addition, recent *in vitro* and *in vivo* studies have suggested that PFAS exposure can induce oxidative stress. An *in vitro* study has revealed that PFAS such as PFOA, PFNA, PFDA, PFUnDA, PFHxS, and PFOS can generate reactive oxygen species and that some of these compounds can reduce the antioxidant capacity in human hepatoma cell line (Wielsøe et al., 2015). Also, multiple *in vivo* studies have demonstrated the potential of PFAS to induce oxidative stress, which provides evidence as an important mechanism in various PFAS-associated health outcomes (Bonato et al., 2020). Furthermore, an animal experiment has identified accumulation of PFAS in eyes (Chang et al., 2012), which suggests its possible contribution to oxidative stress in retina, particularly retinal pigment epithelial cells (Harju, 2022), consequently leading to the development of AMD.

Second, cardiometabolic diseases might mediate a pathway linking PFAS exposure to retina health because previous epidemiological studies have shown associations of PFAS exposure with cardiometabolic diseases, including hypercholesterolemia (Winquist and Steenland, 2014), hypertension (Ding et al., 2022), and diabetes (Gui et al., 2023). These diseases have been reported to be risk factors for AMD (Gemmy Cheung et al., 2013; Ghaem Maralani et al., 2015). However, it is worth noting that associations between PFAS exposure and AMD remain consistent even after adjusting for hypertension, diabetes, and/or total cholesterol.

Third, protective effects of estrogen on AMD might be another mechanism underlying our observations, although evidence is not strong in previous studies. Estrogen is known to play a protective role in the pathogenesis of AMD through its antioxidative and anti-inflammatory effects (Kaarniranta et al., 2015). This protective role of estrogen has been demonstrated in pre- and postmenopausal women as well as rodents through *in vivo* experiments by previous studies (Cascio et al., 2015; Kaarniranta et al., 2015). In addition, PFAS may exert anti-estrogenic or anti-androgenic effects by interacting with hormone receptors (Li et al., 2020) and altering hormone synthesis(Kang et al., 2016) and metabolism (Liu et al., 2023). Nevertheless, the secondary effect of sexual hormone disruption caused by PFAS on AMD needs further investigations.

Inverted U-shaped relationships observed between PFAS exposure and AMD were in line with previous studies reporting similar relationships between PFAS exposure and other health outcomes, including type 2 diabetes (Mancini et al., 2018), cognitive decline (Park et al., 2021), and attention-deficit/hyperactivity disorder symptoms (Kim et al., 2023). One possible explanation is that healthy subjects have higher opportunities of PFAS exposure. Individuals who have frequent recreational outdoor activities such as hiking, camping, skiing, fishing, and bicycle riding have higher chance of PFAS exposure due to the use of PFAS-containing sportswears (Glüge et al., 2020). Another possibility includes a confounding by physical exercise, which plays a protective role in AMD development (Mauschitz et al., 2022). However, we could not rule out the possibility of an actual non-monotonic relationship of endocrine disrupting chemicals (Lagarde et al., 2015). Several hypotheses have been proposed to explain such a non-monotonic relationship, including induction of opposite effects at different dose ranges (e.g., agonism vs. antagonism), negative feedback mechanisms, receptor desensitization, contrasting effects between parent compounds and their metabolites, and formation of mixed-ligand dimers of hormone receptors (Lagarde et al., 2015).

Subgroup analyses revealed effect modification by race/ethnicity, sex, age, and seafood consumption. The stronger association in Non-Hispanic Whites than in others could be attributed to their lower levels of ocular melanin, which could play a role as an antioxidant in preventing AMD (Hu et al., 2008). This finding is in line with a previous study reporting a stronger association between urinary cadmium levels and AMD in Non-Hispanic Whites (Wu et al., 2014). A sex-dependent association between PFAS exposure and AMD is also plausible in that previous epidemiological studies of PFAS exposure have shown male-specific associations with sex hormone disruption and hypercholesterolemia (Winquist and Steenland, 2014; Xie et al., 2021), which can be potential mediating pathways of PFAS-induced AMD development. Differences in baseline levels of sex hormones might be a reason for the more pronounced association of PFAS with AMD in men. We also observed a stronger association in middle-aged adults than in older adults. Given that aging is a critical risk factor for AMD, it is possible that effects of PFAS in older adults might be difficult to observe because they were masked by much stronger effects of aging. Finally, a stronger association in individuals with high seafood consumption was observed. This may be explained by a potential additive interaction between PFAS exposure and heavy metals exposure because seafood consumption can lead to exposure to heavy metals such as cadmium, which is a potential risk factor for AMD (Wu et al., 2014). However, additional adjustment for cadmium did not change the association (Supplemental Table S4).

Limitations should be considered in the interpretation of results. First, our study design was cross-sectional, which might not allow us to determine a causality between serum PFAS concentrations and AMD. Particularly, as discussed earlier, our observations might be affected by behaviors related to PFAS exposure which could be influenced by ocular status. However, this possibility is likely to be minuscule given long elimination half-lives of serum PFAS (estimated to range from 2.1 to 35 years) (ATSDR, 2021). Second, the sample size of this study was not large enough, resulting in low statistical power, which could contribute to the lack of significant associations observed for PFOA and PFHxS. Lastly, although multiple factors were adjusted in our statistical models, we could not rule out residual confounding by unmeasured or unknown factors.

This study also has several strengths. First, this study used a nationally representative population, which allowed us to generalize our findings. Second, the exposure was estimated using reliable biomarkers, i.e., serum concentrations of PFAS, which could minimize misclassification in the exposure assessment. Lastly, we adopted a novel approach of an integrated index representing overall PFAS burden because individual PFAS share common exposure sources and toxicological mechanisms.

## 5. Conclusions

Results of our study provide evidence of the potential contribution of exposure to PFAS, particularly PFOS and PFNA, to AMD risk among adults aged 40 years and older. These results raise the need to reduce current PFAS exposure in order to promote ocular health in middle-aged and older population.

## Data Availability

All data produced are available online at https://www.cdc.gov/nchs/nhanes/index.htm.

