## Supplemental materials for "Exposure to per- and polyfluoroalkyl substances and age-related macular degeneration in U.S. middle-aged and older adults"

Yoon-Hyeong Choi

School of Health and Environmental Science, Korea University,

Anam-ro 145, Seongbuk-gu, Seoul 02841, Korea

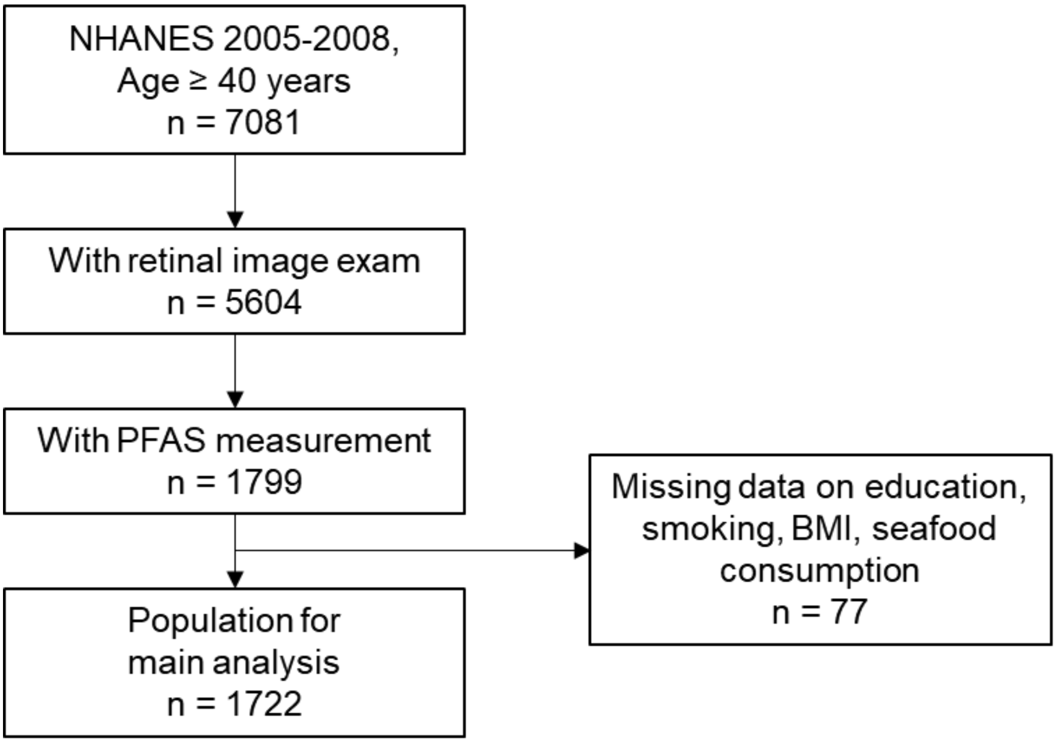


Figure S1. Flow chart showing the selection of subjects used for analysis in this study.

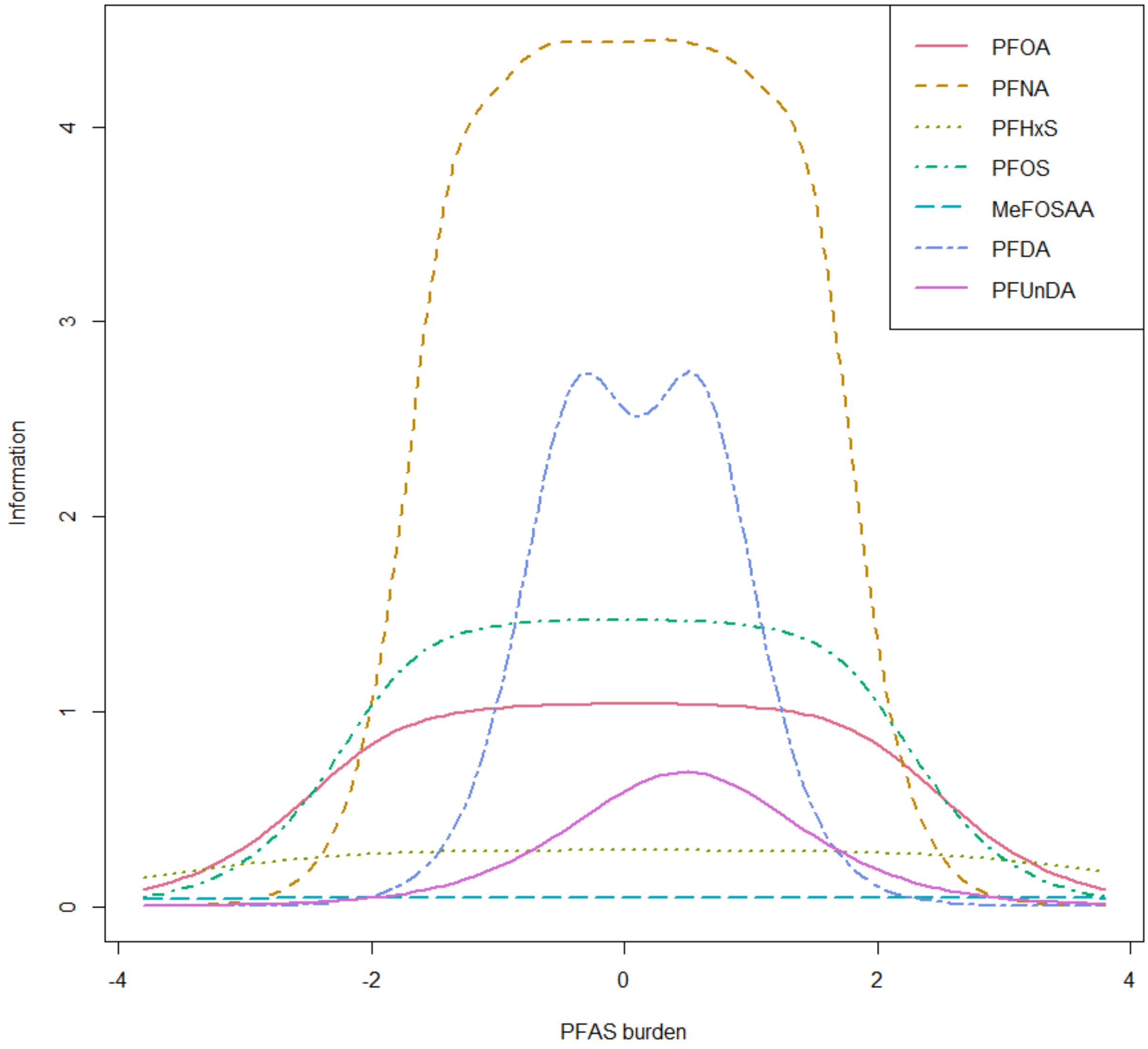


Figure S2. Item information curves of the PFAS item response theory model. The height of each curve at a specific PFAS burden reflects the relative contribution of the corresponding PFAS compound to that PFAS burden level.

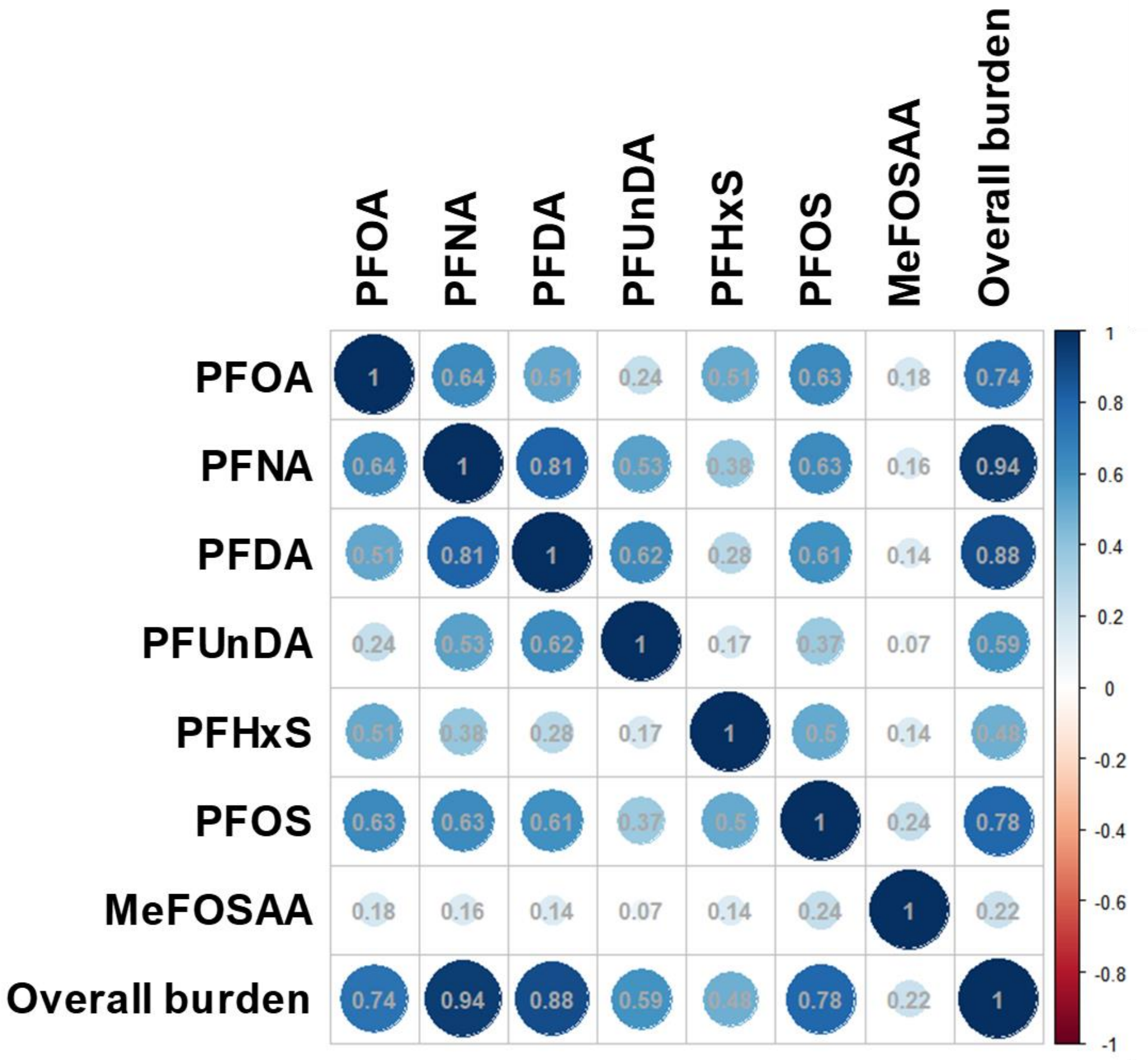


Figure S3. Spearman's correlation coefficients between per- and poly-fluoroalkyl substances (PFAS) concentrations.

Table S1. Target per- and poly-fluoroalkyl substances (PFAS) in this study and their limits of detection (LODs) in the National Health and Nutrition Examination Survey (NHANES) 2005–2008

| PFAS | Abbreviation | Limit of detection (mg/nL) | |
|---|---|---|---|
| | | NHANES 2005–2006 | NHANES 2007–2008 |
| Perfluorheptanoic acid | PFHpA | 0.4 | 0.4 |
| Perfluorooctanoic acid | PFOA | 0.1 | 0.1 |
| Perfluorononanoic acid | PFNA | 0.1 | 0.082 |
| Perfluorodecanoic acid | PFDA | 0.2 | 0.2 |
| Perfluoroundecanoic acid | PFUnDA | 0.2 | 0.2 |
| Perflurododecanoic acid | PFDoDA | 0.2 | 0.2 |
| Perfluorobutane sulfonic acid | PFBS | 0.1 | 0.1 |
| Perfluorohexane sulfonic acid | PFHxS | 0.1 | 0.1 |

| | | | |
|---|---|---|---|
| Perfluorooctane sulfonic acid | PFOS | 0.2 | 0.2 |
| 2-(N-Methyl-perfluorooctane sulfonamido) acetic acid | MeFOSAA | 0.2 | 0.2 |
| 2-(N-Ethyl-perfluorooctane sulfonamido) acetic acid | EtFOSAA | 0.2 | 0.2 |
| Perfluorooctane sulfonamide | PFOSA | 0.1 | 0.1 |

Table S2. Number of participants by Perfluorononanoic acid (PFNA), Perfluorooctane sulfonic acid (PFOS), and/or overall per- and poly-fluoroalkyl substances (PFAS) burden classification

| | | PFNA | | | PFOS | | | Total |
|---|---|---|---|---|---|---|---|---|
| | | Low | Medium | High | Low | Medium | High | |
| Overall PFAS burden | Low | 498 | 74 | 2 | 417 | 141 | 16 | 574 |
| | Medium | 87 | 422 | 65 | 139 | 298 | 137 | 574 |
| | High | 1 | 89 | 484 | 23 | 130 | 421 | 574 |
| Total | | 586 | 585 | 551 | 579 | 569 | 574 | 1722 |

Table S3. Survey-weighted odds ratios (ORs) and 95% confidence intervals (CIs) for any age-related macular degeneration (AMD) by serum PFAS levels with additional adjustment for hypertension, diabetes, and/or total cholesterol

| | Model 2 in main text (n=1,722) | + hypertension (n=1,704) | + diabetes (n=1,720) | + total cholesterol (n=1,722) | + hypertension, diabetes, and total cholesterol (n=1,702) |
|---|---|---|---|---|---|
| | OR (95% CI) | OR (95% CI) | OR (95% CI) | OR (95% CI) | OR (95% CI) |
| PFOA | | | | | |
| Low | 1 (reference) | 1 (reference) | 1 (reference) | 1 (reference) | 1 (reference) |
| Medium | 1.32 (0.81, 2.14) | 1.35 (0.81, 2.24) | 1.29 (0.80, 2.10) | 1.30 (0.81, 2.09) | 1.31 (0.79, 2.16) |
| High | 1.40 (0.78, 2.52) | 1.44 (0.79, 2.60) | 1.40 (0.78, 2.51) | 1.36 (0.74, 2.49) | 1.40 (0.76, 2.55) |
| *p* for linear trend | 0.29 | 0.26 | 0.29 | 0.35 | 0.31 |
| PFNA | | | | | |
| Low | 1 (reference) | 1 (reference) | 1 (reference) | 1 (reference) | 1 (reference) |
| Medium | 1.74 (0.98, 3.09) | 1.78 (0.99, 3.20) | 1.71 (0.97, 3.02) | 1.72 (0.97, 3.05) | 1.73 (0.97, 3.09) |
| High | 1.46 (0.77, 2.76) | 1.42 (0.73, 2.75) | 1.46 (0.77, 2.79) | 1.45 (0.77, 2.74) | 1.41 (0.73, 2.74) |
| *p* for linear trend | 0.28 | 0.33 | 0.28 | 0.28 | 0.34 |
| PFHxS | | | | | |
| Low | 1 (reference) | 1 (reference) | 1 (reference) | 1 (reference) | 1 (reference) |
| Medium | 1.61 (0.80, 3.25) | 1.63 (0.78, 3.38) | 1.58 (0.76, 3.27) | 1.60 (0.80, 3.23) | 1.61 (0.76, 3.40) |
| High | 1.36 (0.81, 2.29) | 1.40 (0.83, 2.37) | 1.35 (0.79, 2.29) | 1.32 (0.78, 2.25) | 1.38 (0.81, 2.34) |
| *p* for linear trend | 0.28 | 0.22 | 0.29 | 0.33 | 0.27 |
| PFOS | | | | | |
| Low | 1 (reference) | 1 (reference) | 1 (reference) | 1 (reference) | 1 (reference) |
| Medium | 1.49 (0.71, 3.15) | 1.48 (0.69, 3.14) | 1.49 (0.71, 3.13) | 1.48 (0.70, 3.12) | 1.44 (0.67, 3.09) |
| High | 1.99 (1.05, 3.79) | 2.04 (1.06, 3.93) | 1.96 (1.03, 3.72) | 1.95 (1.02, 3.71) | 1.97 (1.02, 3.78) |
| *p* for linear trend | 0.03 | 0.03 | 0.04 | 0.04 | 0.04 |
| Overall PFAS burden (IRT score) | | | | | |
| Low | 1 (reference) | 1 (reference) | 1 (reference) | 1 (reference) | 1 (reference) |
| Medium | 2.18 (1.18, 4.04) | 2.22 (1.17, 4.21) | 2.18 (1.18, 4.04) | 2.14 (1.16, 3.96) | 2.18 (1.14, 4.16) |
| High | 1.88 (0.96, 3.68) | 1.88 (0.94, 3.73) | 1.85 (0.95, 3.62) | 1.86 (0.95, 3.65) | 1.83 (0.92, 3.64) |
| *p* for linear trend | 0.10 | 0.11 | 0.11 | 0.11 | 0.13 |

Note: Perfluorooctanoic acid, PFOA; perfluorononanoic acid, PFNA; perfluorooctane sulfonic acid, PFOS; per- and poly-fluoroalkyl substances, PFAS.

Table S4. Survey-weighted odds ratios (ORs) and 95% confidence intervals (CIs) for any age-related macular degeneration (AMD) by serum PFAS levels with additional adjustment for blood cadmium concentrations

| | Model 2 in main text (n=1,722) | + blood cadmium[a] (n=1,719) |
|---|---|---|
| | OR (95% CI) | OR (95% CI) |
| PFOA | | |
| Low | 1 (reference) | 1 (reference) |
| Medium | 1.32 (0.81, 2.14) | 1.36 (0.82, 2.25) |
| High | 1.40 (0.78, 2.52) | 1.41 (0.76, 2.60) |
| *p* for linear trend | 0.29 | 0.31 |
| PFNA | | |
| Low | 1 (reference) | 1 (reference) |
| Medium | 1.74 (0.98, 3.09) | 1.73 (0.98, 3.04) |
| High | 1.46 (0.77, 2.76) | 1.46 (0.77, 2.75) |
| *p* for linear trend | 0.28 | 0.28 |
| PFHxS | | |
| Low | 1 (reference) | 1 (reference) |
| Medium | 1.61 (0.80, 3.25) | 1.61 (0.81, 3.23) |
| High | 1.36 (0.81, 2.29) | 1.38 (0.82, 2.34) |
| *p* for linear trend | 0.28 | 0.25 |
| PFOS | | |
| Low | 1 (reference) | 1 (reference) |
| Medium | 1.49 (0.71, 3.15) | 1.47 (0.71, 3.07) |
| High | 1.99 (1.05, 3.79) | 2.01 (1.04, 3.86) |
| *p* for linear trend | 0.03 | 0.04 |
| Overall PFAS burden (IRT score) | | |
| Low | 1 (reference) | 1 (reference) |
| Medium | 2.18 (1.18, 4.04) | 2.19 (1.16, 4.12) |
| High | 1.88 (0.96, 3.68) | 1.91 (0.98, 3.75) |
| *p* for linear trend | 0.10 | 0.09 |

Note: Perfluorooctanoic acid, PFOA; perfluorononanoic acid, PFNA; perfluorooctane sulfonic acid, PFOS; per- and poly-fluoroalkyl substances, PFAS.
[a] Quartiles of blood cadmium concentrations were additionally adjusted for based on previous literature (Wu et al., 2014).
